# Analytical and clinical evaluation of four anti-SARS-CoV-2 serologic (IgM, IgG, and total) immunoassays

**DOI:** 10.1101/2020.10.23.20217810

**Authors:** Victoria Higgins, Anselmo Fabros, Xiao Yan Wang, Maria Bhandari, David J. Daghfal, Vathany Kulasingam

## Abstract

**Introduction:** Coronavirus disease 2019 (COVID-19), caused by severe acute respiratory syndrome coronavirus 2 (SARS-CoV-2), is diagnosed by molecular-based detection of SARS-CoV-2 RNA. Serologic testing detects antibodies specific to SARS-CoV-2 and IgM specifically may serve as an adjunct test to PCR early in disease. We evaluated the Abbott anti-SARS-CoV-2 IgM and IgG assays along with DiaSorin anti-SARS-CoV-2 IgG and Roche anti-SARS-CoV-2 Total.

**Methods:** Specimens from 175 PCR-positive patients and 107 control specimens were analyzed using Abbott IgM and IgG, DiaSorin IgG, and Roche Total (IgA, IgG, IgM) assays. Sensitivity, specificity, cross-reactivity, concordance between assays, trends over time, positive predictive value (PPV), and negative predictive value (NPV) were determined.

**Results:** Abbott IgM sensitivity was 63.6% at 0 days post-PCR positivity, 76.5% at 1-5d, 76.3% at 6-14d, 85.2% at 15-30d, and 63.6% at >30d. All assays exhibited highest sensitivity 15-30d post-PCR positivity (83.3-85.2%). Combining Abbott IgM and IgG improved sensitivity by 22.7% compared to IgG alone when tested 0d post-PCR positivity. All assays had a specificity of 100% and only Abbott IgG exhibited cross-reactivity (anti-dsDNA). Cohen’s kappa varied between 0.86-0.93. Time to seroconversion from PCR positivity was lowest for Abbott IgM and highest for Abbott IgG. NPV was highest for Abbott IgM <14 days post-PCR positivity and Abbott IgG ≥14 days.

**Conclusion:** The Abbott IgM assay exhibited the earliest response and greatest signal in most patients evaluated for serial sampling and had the highest NPV <14 days post-PCR positivity, suggesting its potential utility as an adjunct test to PCR early in disease course.

## Introduction

Coronavirus disease 2019 (COVID-19), caused by the severe acute respiratory syndrome coronavirus 2 (SARS-CoV-2), emerged from Wuhan, China in late 2019 (1). COVID-19 was first declared a Public Health Emergency of International Concern in January 2020 by the World Health Organization (WHO) and has infected over 37 million people globally, causing over one million deaths as of 11 October 2020 (2). Clinical manifestations of COVID-19 illness vary in severity between patients from asymptomatic to severe pneumonia, acute respiratory distress syndrome, sepsis, and/or multisystem organ failure (3). COVID-19 is diagnosed by molecular-based detection of SARS-CoV-2 RNA, most commonly by reverse transcription-polymerase chain reaction (RT-PCR) in nasopharyngeal and/or oropharyngeal specimens (4). Viral RNA can be detected in these specimens as early as the first day of symptom onset, peaks within the first week, and can remain positive beyond three weeks in severe cases (5,6). However, PCR positivity only reflects viral RNA detection, not necessarily the presence of viable virus (7), and its predictive value varies with time from exposure and symptom onset (8). For example, one study reported the probability of a false negative result to be 100% on day 1 of infection, 67% on day 4, 38% on day 5 (symptom onset), 20% on day 8, and 66% on day 21 (8).

Serologic testing detects antibodies (e.g. IgG, IgM) specific to SARS-CoV-2 in blood, serum, or plasma. While serologic testing is not useful on its own for COVID-19 diagnosis (9), it may serve as an adjunct to molecular-based testing for COVID-19 diagnosis if used ≥15 days after symptom onset in cases with suggestive clinical presentation, but where RT-PCR results are negative or not available (9). As IgM is a marker of acute infection, it may be a useful tool to combine with PCR to improve sensitivity and specificity early in the disease course (i.e. <14 days after symptom onset) (10–12). Antibody response has been reported to correlate with disease severity, with more severe cases exhibiting immediate seroconversion (13). Furthermore, antibody titers were found to be higher in severe compared to non-severe cases two weeks post-symptom onset (12). Serologic testing may also have clinical utility for surveying asymptomatic infection in close contacts and population-level assessment of the prevalence of past SARS-CoV-2 infection (12,14,15). While data are still limited, there is mounting evidence that antibodies detected by commercial serologic assays correlate with neutralization capacity (16) and confer some resistance to re-infection (17,18). Thus, serologic testing may also have clinical utility for international travel authorization, assessing reinfection risk in workplaces, and facilitating economic activity resumption.

In order to demonstrate an adequate positive predictive value, it is important for serologic assays to demonstrate high sensitivity and specificity, particularly when seroprevalence is low (19). It has been suggested that laboratories should implement SARS-CoV-2 serologic tests that have manufacturer-claimed sensitivity ≥95% and specificity ≥99.5% based on specimens obtained ≥14 days after symptom onset or PCR positivity (20). We evaluated the sensitivity, specificity, cross-reactivity, concordance, trends over time, positive predictive value, and negative predictive value for four serologic assays: Abbott anti-SARS-CoV-2 IgM serologic assay, Abbott anti-SARS-CoV-2 IgG, DiaSorin anti-SARS-CoV-2 IgG, and Roche anti-SARS-CoV-2 Total assays.

## Materials and Methods

### Sample Collection and Analysis

This work was exempt from Quality Improvement (QI) review and Research Ethics Board (REB) approval at the University Health Network (UHN; Toronto, Canada). Presence or absence of SARS-CoV-2 infections was determined by SARS-CoV-2 viral RNA detection in nasopharyngeal swabs tested at UHN microbiology lab on assays validated for clinical use (Seegene Allplex 2019-nCoV assay which has been approved by Health Canada for Emergency Use Authorization and verified by UHM microbiology lab). Deidentified residual patient serum and plasma samples were collected from UHN and analyzed using four anti-SARS-CoV-2 serologic assays at UHN, including SARS-CoV-2 IgG and SARS-CoV-2 IgM on the Abbott ARCHITECT® i (Abbott Diagnostics), SARS-CoV-2 S1/S2 IgG on the LIAISON® XL (DiaSorin), and Elecsys® anti-SARS-CoV-2 total (IgA, IgG, IgM) on the cobas e411 (Roche Diagnostics). Details and performance characteristics of these four serologic assays are described in **Table S1**. The Abbott anti-SARS-CoV-2 IgG and IgM assays are qualitative chemiluminescent microparticle immunoassays (CMIA). The anti-SARS-CoV-2 IgG assay detects IgG antibodies to the nucleocapsid protein of SARS-CoV-2, while the anti-SARS-CoV-2 IgM assay detects IgM antibodies to the receptor-binding domain (RBD) of the spike protein (S1) of SARS-CoV-2. The DiaSorin anti-SARS-CoV-2 S1/S2 IgG assay is a qualitative chemiluminescent immunoassay (CLIA) that detects IgG antibodies against the spike protein (S1 and S2 subunits) of SARS-CoV-2. Lastly, the Roche anti-SARS-CoV-2 total assay is a qualitative electrochemiluminescence immunoassay (ECLIA) that detects IgA, IgM and IgG antibodies to the nucleocapsid protein of SARS-CoV-2. Preventative maintenance, function checks, calibration, and internal quality control were performed according to the manufacturer’s instructions.

### Sensitivity

To determine the sensitivity of four anti-SARS-CoV-2 antibody assays, serum or plasma samples were collected from 175 patients that were confirmed positive for SARS-CoV-2 infection by PCR testing within the previous 0-73 days. Sensitivity was calculated as true positive / (true positive + false negative), where true positivity was defined as PCR positivity. Total sensitivity and sensitivity for various categories of days post-PCR positivity were determined.

### Cross-reactivity & specificity

To determine the cross-reactivity of the four anti-SARS-CoV-2 antibody assays, serum or plasma samples were collected from 107 patients that were positive for viruses other than SARS-CoV-2 (e.g. hepatitis A, hepatitis B, hepatitis C, human immunodeficiency virus, rubella, Epstein-Barr virus, cytomegalovirus, respiratory syncytial virus, enterovirus, rhinovirus, influenza A, influenza B, metapneumovirus, BK virus), had autoantibodies or a known autoimmune condition (e.g. anti-dsDNA, rheumatoid factor, anti-centromere, anti-SSA, anti-Sm, anti-SmRNP, anti-RiboP, celiac disease, anti-MPO, anti-PR3, anti-CCP, antinuclear antibodies), had elevations of other analytes (e.g. creatinine, C-reactive protein, IgA, IgG, IgM), or had the influenza vaccine in 2019. The percentage of samples that were incorrectly identified as positive for SARS-CoV-2 antibodies was determined for each assay.

Specificity was assessed using 52 of these samples that were collected from patients in 2019, before SARS-CoV-2 was thought to be circulating in Ontario, Canada (n=32), or were confirmed to be SARS-CoV-2 negative by PCR (n=20). Specificity was calculated as true negative / (true negative + false positive), where true negativity was defined as negative by PCR or sample collection prior to the circulation of SARS-CoV-2.

### Concordance

Concordance between the four anti-SARS-CoV-2 antibody assays was determined using all 175 samples from the sensitivity analysis and all 107 samples from the cross-reactivity and specificity analysis. Overall percent agreement, positive percent agreement (PPA), negative percent agreement (NPA), and Cohen’s kappa were calculated for each pair-wise comparison.

### Serial sampling to model antibody response

Serial serum and plasma samples (n=6-20) were collected from five patients that were positive for SARS-CoV-2 by PCR. Data was expressed as a ratio of the response (i.e. assay signal) to the positivity cut-off of the assay. This ratio was examined over time since PCR positivity.

### Positive predictive value and negative predictive value

Sensitivity for <14 days and ≥14 days post-PCR positivity, as well as specificity for each of the four anti-SARS-CoV-2 serologic assays as determined in this study were used to estimate the positive predictive value (PPV) and negative predictive value (NPV) at seroprevalence values of 1%, 5%, and 10%.

## Results

### Sensitivity

**Fig. 1** shows anti-SARS-CoV-2 antibody results for four SARS-CoV-2 serologic assays plotted against days since PCR positivity. Overall, the assay signal does not correlate with days since PCR positivity, with a wide range of signal results observed across all days since PCR positivity.

**Fig. 1:**
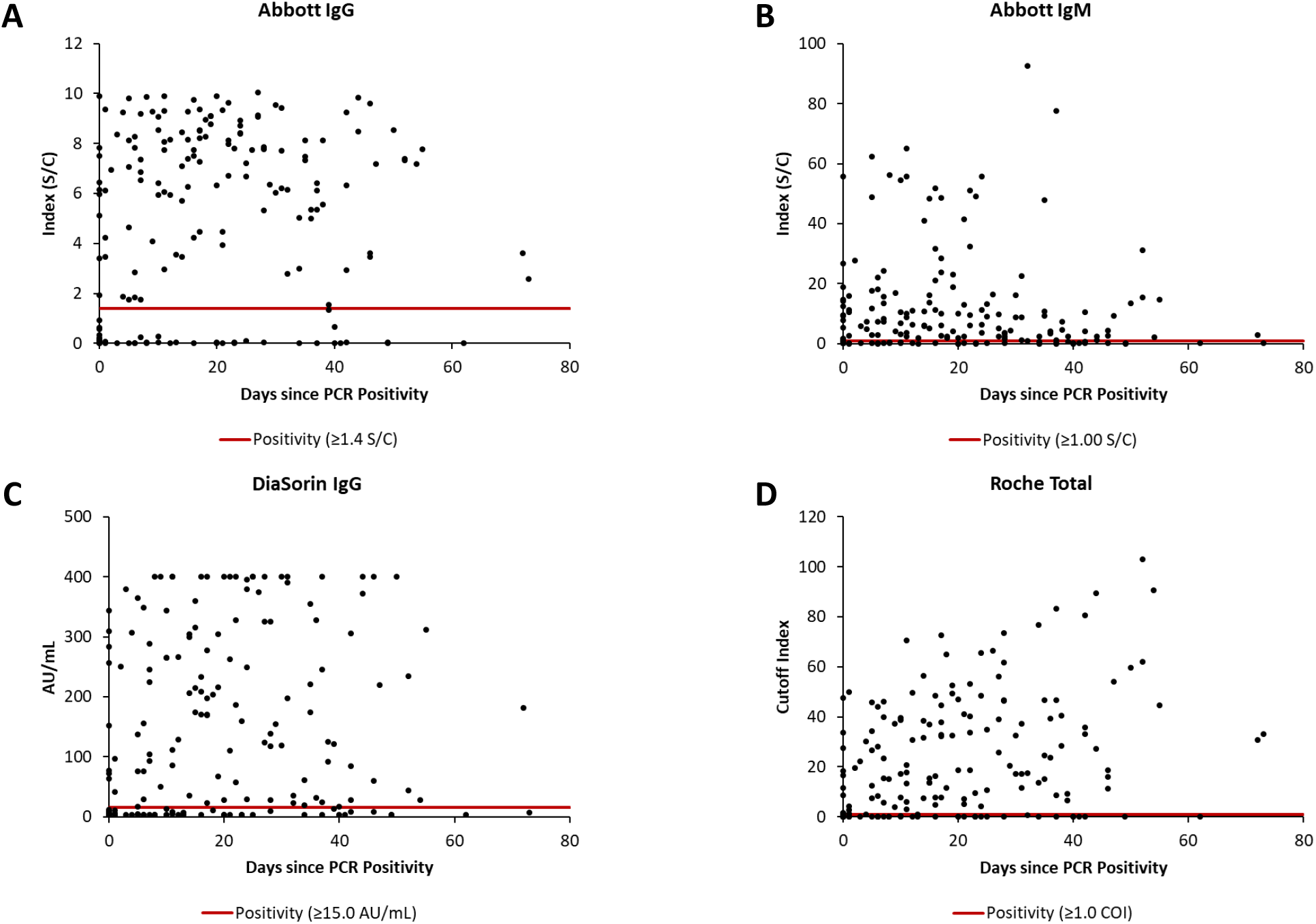
Anti-SARS-CoV-2 antibody results plotted against days since PCR positivity for four SARS-CoV-2 serologic assays: **(A)** Abbott anti-SARS-CoV-2 IgG, **(B)** Abbott anti-SARS-CoV-2 IgM, **(C)** DiaSorin anti-SARS-CoV-2 IgG, and **(D)** Roche anti-SARS-CoV-2 total. The solid red horizontal line indicates the assay specific positivity cut-off.

Anti-SARS-CoV-2 antibody sensitivity of four SARS-CoV-2 serologic assays is summarized in **Table 1** and **Fig. 2**. The total sensitivity (independent of days since PCR positivity) ranged from 69.0% to 74.9% across assays. All assays exhibited greater sensitivity ≥14 days post-PCR positivity (76.5%-82.4%) compared to <14 days (52.1%-64.4%), although Abbott anti-SARS-CoV-2 IgM exhibited the greatest sensitivity among the assays <14 days. All assays exhibited the highest sensitivity between 15-30 days post-PCR positivity, ranging from 83.3% to 85.2%,. As expected, the lowest sensitivity was observed for samples collected the same day as PCR positivity, ranging from 38.1% to 63.6% with the highest sensitivity observed for Abbott anti-SARS-CoV-2 IgM. Overall, 20.6% of patients positive for SARS-CoV-2 by PCR were negative across all four serologic assays, in-line with the large immunocompromised population at UHN.

**Table 1.** Sensitivity comparison among four SARS-CoV-2 serologic assays by days post-PCR positivity

|  |  | Days Post-PCR Positivity |  |  |  |  |  |  |
| --- | --- | --- | --- | --- | --- | --- | --- | --- |
|  | Total | 0 | 1-5 | 6-14 | 15-30 | >30 | <14 | ≥14 |
| **n** | 175 | 22 | 17 | 38 | 54 | 44 | 73 | 102 |
| **Abbott IgG** | 74.9% | 40.9% | 76.5% | 76.3% | 85.2% | 77.3% | 64.4% | 82.4% |
| **Abbott IgM** | 74.3% | 63.6% | 76.5% | 76.3% | 85.2% | 63.6% | 71.2% | 76.5% |
| **DiaSorin IgG<sup>a</sup>** | 69.0% | 38.1% | 52.9% | 65.8% | 83.3% | 75.0% | 52.1% | 80.4% |
| **Roche Total<sup>b</sup>** | 73.6% | 45.5% | 70.6% | 71.1% | 83.3% | 79.1% | 61.6% | 81.4% |

\*Sensitivity calculated as true positive / (true positive + false negative), with a true positive determine by PCR positivity
<sup>a</sup>Sample size for DiaSorin IgG was 174. (n=72 for Day <14; n=21 for Day 0)
<sup>b</sup>Sample size for Roche Total was 174. (n=101 for Day ≥14; n=43 for Day >30)

**Fig. 2:**
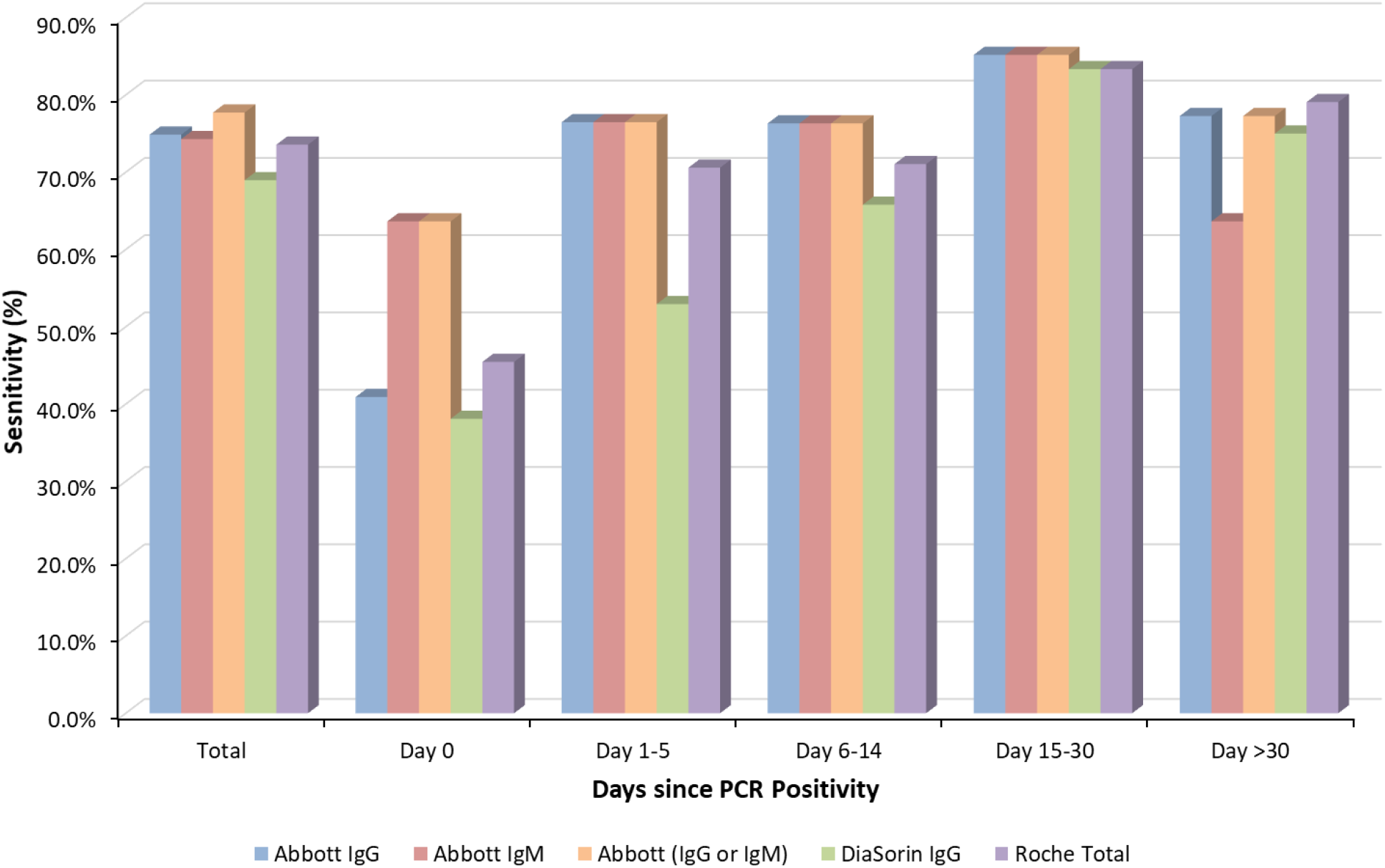
Total sensitivity and sensitivity for 0, 1-5, 6-14, 15-30, and >30 days since PCR positivity for four Abbott anti-SARS-CoV-2 IgG, Abbott anti-SARS-CoV-2 IgM, Abbott anti-SARS-CoV-2 IgG and IgM together (either assay is positive), DiaSorin anti-SARS-CoV-2 IgG, and Roche anti-SARS-CoV-2 total

When including both Abbott anti-SARS-CoV-2 IgG and IgM assays, total sensitivity improved from 74.9% (IgG alone) and 74.3% (IgM alone) to 77.7%, as shown in **Table 2** and **Fig. 2**. IgG measurement improved sensitivity for samples >30 days after PCR positivity (13.6% were positive for IgG only). On the other hand, IgM measurement improved sensitivity for samples collected the same day as PCR positivity (22.7% were positive for IgM only).

**Table 2.** Sensitivity comparison among Abbott IgM and Abbott IgG anti-SARS-CoV-2 serologic assays by days post-PCR positivity

|  |  | Days Post-PCR Positivity |  |  |  |  |  |  |
| --- | --- | --- | --- | --- | --- | --- | --- | --- |
|  | Total | 0 | 1-5 | 6-14 | 15-30 | >30 | <14 | ≥14 |
| **n** | 175 | 22 | 17 | 38 | 54 | 44 | 73 | 102 |
| **Total Abbott IgG** | 74.9% | 40.9% | 76.5% | 76.3% | 85.2% | 77.3% | 64.4% | 82.4% |
| **Total Abbott IgM** | 74.3% | 63.6% | 76.5% | 76.3% | 85.2% | 63.6% | 71.2% | 76.5% |
| **Abbott IgM and IgG** | 71.4% | 40.9% | 76.5% | 76.3% | 85.2% | 63.6% | 64.4% | 76.5% |
| **Abbott IgM or IgG** | 77.7% | 63.6% | 76.5% | 76.3% | 85.2% | 77.3% | 71.2% | 82.4% |
| **Abbott IgG Only** | 3.4% | 0.0% | 0.0% | 0.0% | 0.0% | 13.6% | 0.0% | 5.9% |
| **Abbott IgM Only** | 2.9% | 22.7% | 0.0% | 0.0% | 0.0% | 0.0% | 6.8% | 0.0% |

\*Sensitivity calculated as true positive / (true positive + false negative), with a true positive determine by PCR positivity

### Cross-reactivity & specificity

Of the 107 control patient samples measured, all were negative for Abbott anti-SARS-CoV-2 IgM, DiaSorin anti-SARS-CoV-2 IgG, and Roche anti-SARS-CoV-2 total. One sample positive for anti-dsDNA antibody (157 IU/mL as measured on the Bioplex 2200 platform) was positive for Abbott anti-SARS-CoV-2 IgG with an index value of 3.49 (positive: ≥1.4). Four additional samples positive for anti-dsDNA antibody (concentrations ranging from 19-44 IU/mL) were negative for anti-SARS-CoV-2 antibodies by all four assays. All 52 control patient samples that were collected pre-COVID-19 or were PCR negative were negative by all four serologic assays. Therefore, all four serologic assays exhibited an assay specificity of 100%.

### Concordance

**Table 3** and **Fig. S1-S2** summarize the concordance between the four serologic assays. The overall percent agreement and Cohen’s kappa varied between 93.2-96.4% and 0.86-0.93, respectively. PPA and NPA varied between 89.1-96.7% and 93.1-99.3%, respectively. The strongest overall percent agreement and Cohen’s kappa were observed between Abbott anti-SARS-CoV-2 IgG and Roche anti-SARS-CoV-2 total, which both target the nucleocapsid protein of SARS-CoV-2. The weakest overall percent agreement and Cohen’s kappa were observed between Abbott anti-SARS-CoV-2 IgM and DiaSorin anti-SARS-CoV-2 IgG. This was the result of a low PPA (89.1%) due to 14 results positive by Abbott anti-SARS-CoV-2 IgM, but negative by DiaSorin anti-SARS-CoV-2 IgG.

**Table 3.**
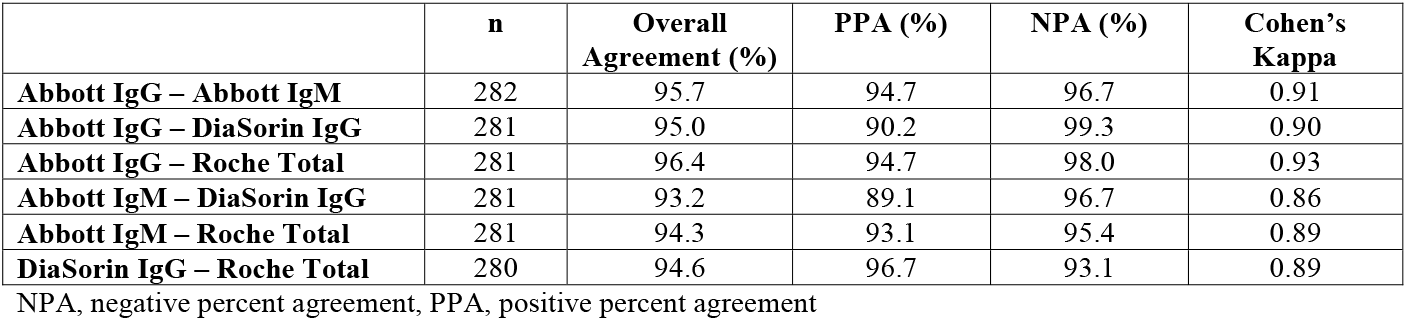
Concordance between four anti-SARS-CoV-2 serologic assays

|  | **n** | **Overall Agreement (%)** | **PPA (%)** | **NPA (%)** | **Cohen's Kappa** |
| --- | --- | --- | --- | --- | --- |
| **Abbott IgG – Abbott IgM** | 282 | 95.7 | 94.7 | 96.7 | 0.91 |
| **Abbott IgG – DiaSorin IgG** | 281 | 95.0 | 90.2 | 99.3 | 0.90 |
| **Abbott IgG – Roche Total** | 281 | 96.4 | 94.7 | 98.0 | 0.93 |
| **Abbott IgM – DiaSorin IgG** | 281 | 93.2 | 89.1 | 96.7 | 0.86 |
| **Abbott IgM – Roche Total** | 281 | 94.3 | 93.1 | 95.4 | 0.89 |
| **DiaSorin IgG – Roche Total** | 280 | 94.6 | 96.7 | 93.1 | 0.89 |

NPA, negative percent agreement, PPA, positive percent agreement

### Serial sampling to model antibody response

Trends in anti-SARS-CoV-2 antibody levels over time since PCR positivity, as measured by four serologic assays, are shown in **Fig. 3-7**. It is evident that the antibody response to SARS-CoV-2 infection varies between individual patients and across serologic assay platforms and isotypes. Patients 1 and 2 (**Fig. 3-4**), exhibited a rapid rise in Abbott anti-SARS-CoV-2 IgM levels, reaching a response 60 times the positivity cut-off for the assay and either plateaued (Patient 1) or decreased (Patient 2) around one week post-PCR positivity. All other assays exhibited a lower response that either gradually increased or plateaued around five days post-PCR positivity. Patient 3 (**Fig. 5**) exhibited the greatest response for Roche anti-SARS-CoV-2 total, reaching over 60 times the positivity cut-off for the assay and plateauing around 35 days post-PCR positivity, while the other assays exhibited a lower response. Patients 4 and 5 (**Fig. 6-7**) exhibited the most rapid response in Abbott anti-SARS-CoV-2 IgM, peaking around day 12 post-PCR positivity and subsequently decreasing. Both patients exhibited a delayed response for Roche anti-SARS-CoV-2 total, which subsequently peaked around 40 times the positivity cut-off between 30-45 days post-PCR positivity.

**Fig. 3:**
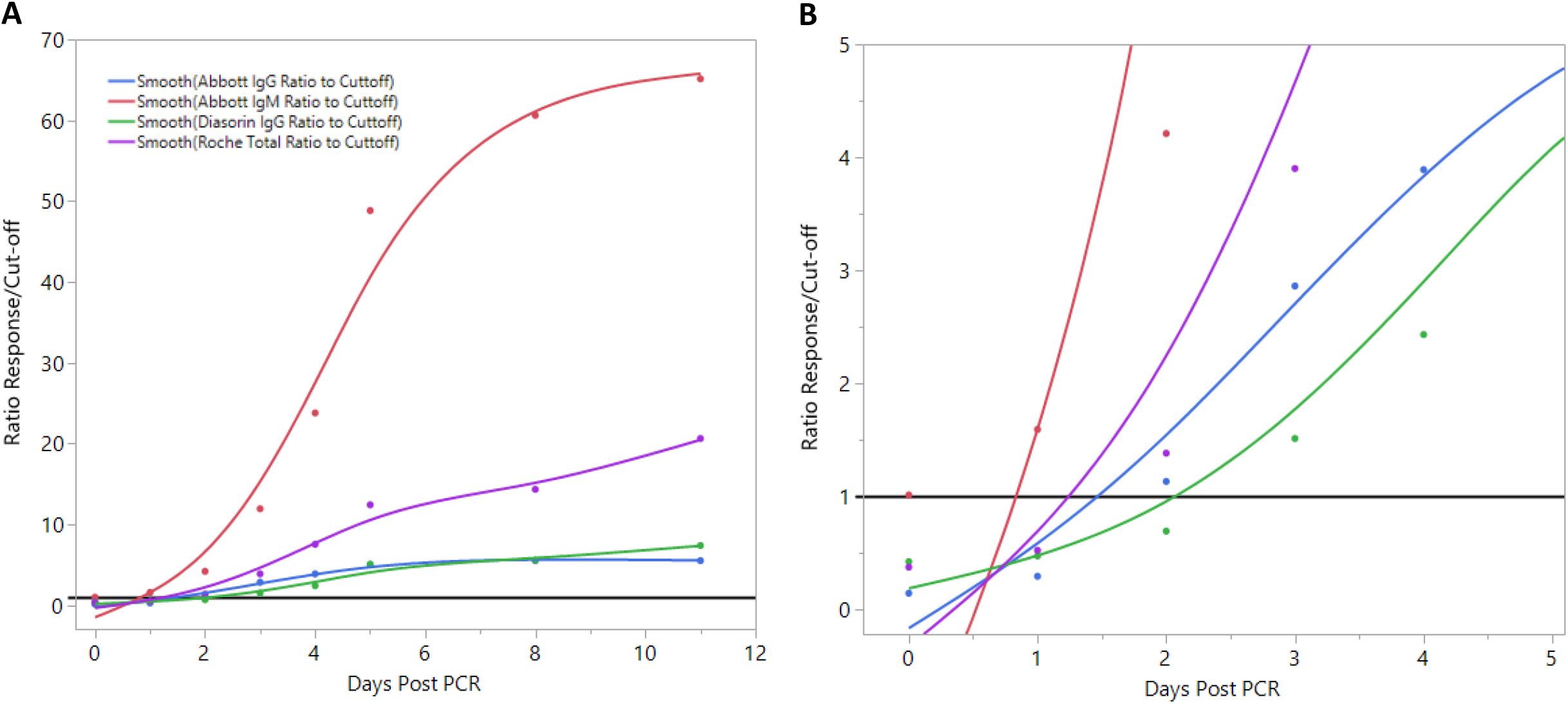
**(A)** Anti-SARS-CoV-2 antibody response by days since PCR positivity in Patient 1 as depicted by serial sampling. Data is plotted as a ratio of the response (i.e. assay signal) to the positivity cut-off of the assay. This ratio is plotted against days since PCR positivity. **(B)** Zoomed in view to better visualize the response around the positivity cut-off (i.e. ratio =1)

**Fig. 4:**
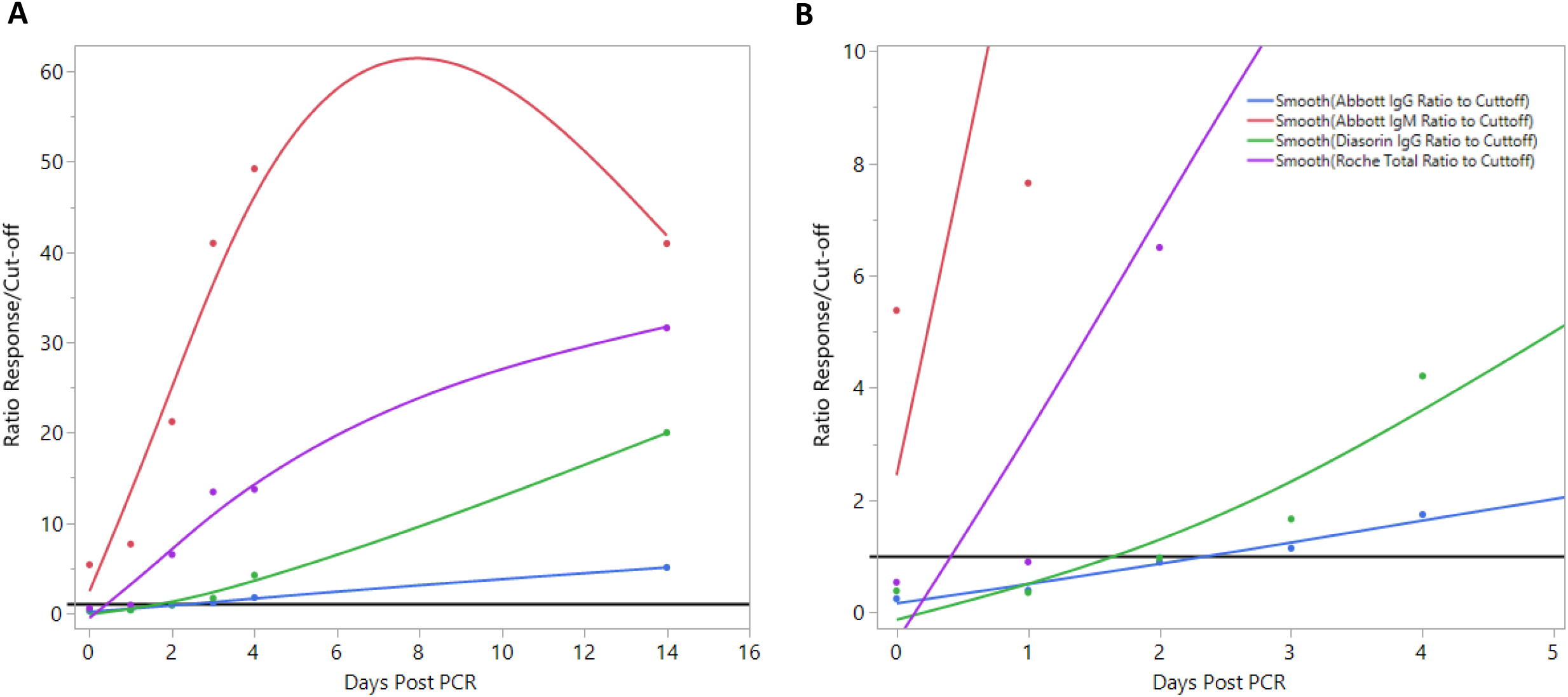
**(A)** Anti-SARS-CoV-2 antibody response by days since PCR positivity in Patient 2 as depicted by serial sampling. Data is plotted as a ratio of the response (i.e. assay signal) to the positivity cut-off of the assay. This ratio is plotted against days since PCR positivity. **(B)** Zoomed in view to better visualize the response around the positivity cut-off (i.e. ratio =1)

**Fig. 5:**
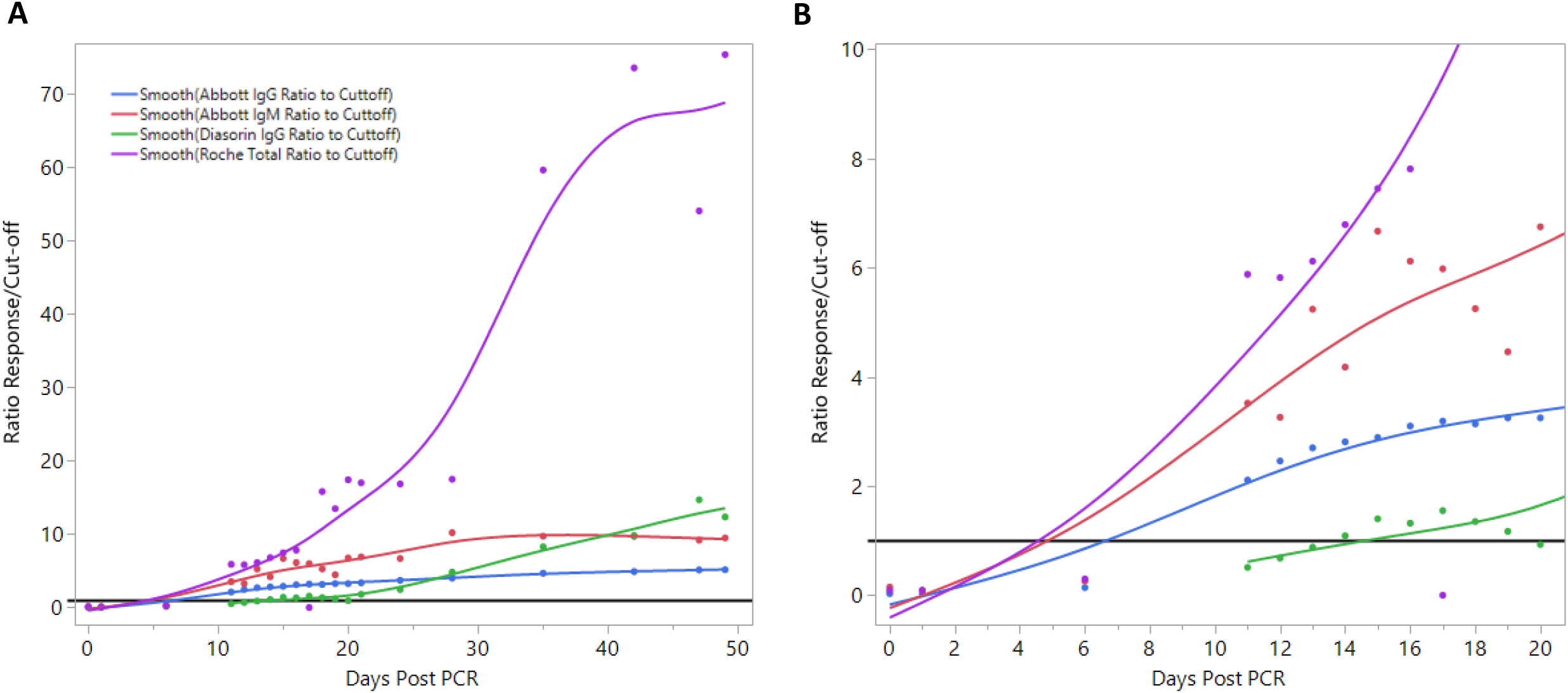
**(A)** Anti-SARS-CoV-2 antibody response by days since PCR positivity in Patient 3 as depicted by serial sampling. Data is plotted as a ratio of the response (i.e. assay signal) to the positivity cut-off of the assay. This ratio is plotted against days since PCR positivity. **(B)** Zoomed in view to better visualize the response around the positivity cut-off (i.e. ratio =1)

**Fig. 6:**
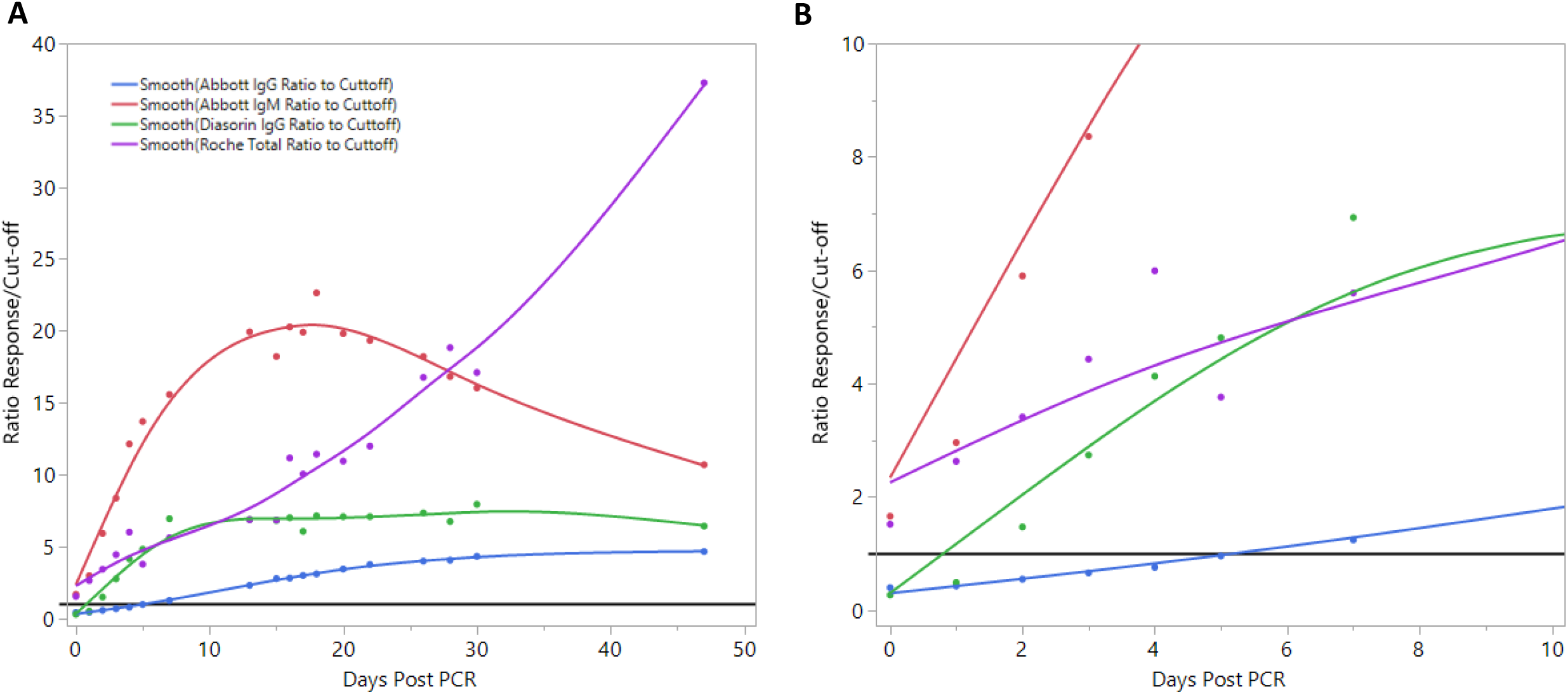
**(A)** Anti-SARS-CoV-2 antibody response by days since PCR positivity in Patient 4 as depicted by serial sampling. Data is plotted as a ratio of the response (i.e. assay signal) to the positivity cut-off of the assay. This ratio is plotted against days since PCR positivity. **(B)** Zoomed in view to better visualize the response around the positivity cut-off (i.e. ratio =1)

**Fig. 7:**
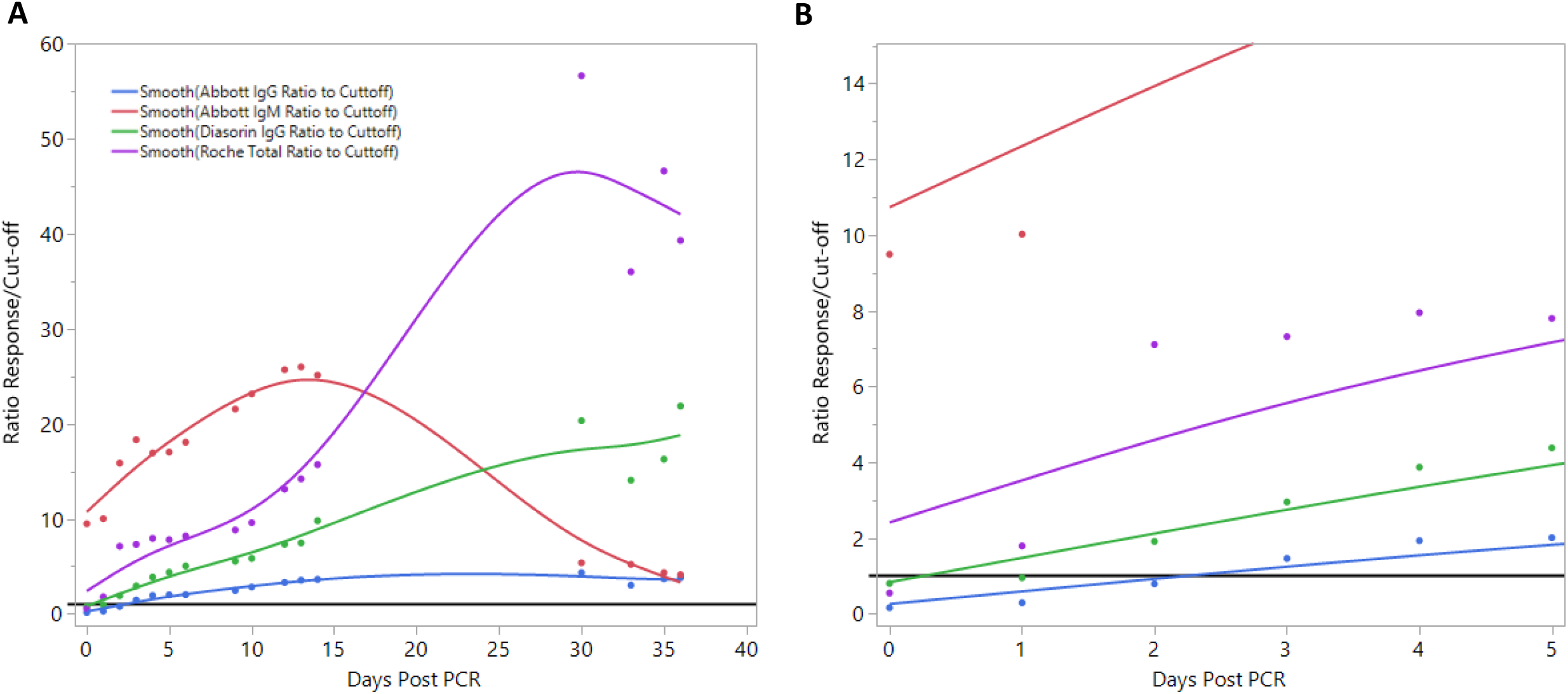
**(A)** Anti-SARS-CoV-2 antibody response by days since PCR positivity in Patient 5 as depicted by serial sampling. Data is plotted as a ratio of the response (i.e. assay signal) to the positivity cut-off of the assay. This ratio is plotted against days since PCR positivity. **(B)** Zoomed in view to better visualize the response around the positivity cut-off (i.e. ratio =1)

**Table 4** summarizes days from PCR positivity to seroconversion as determined for each serologic assay. Overall, the time to seroconversion was lowest for Abbott anti-SARS-CoV-2 IgM (median: 0 days; mean: 2.2 days), followed by Roche anti-SARS-CoV-2 total, which also contains IgM (median: 2 days; mean: 3.2 days). Abbott anti-SARS-CoV-2 IgG (median: 3 days; mean: 5.2 days) and DiaSorin anti-SARS-CoV-2 IgG (median: 3 days; mean: 4.8 days) exhibited longer times to seroconversion.

**Table 4.**
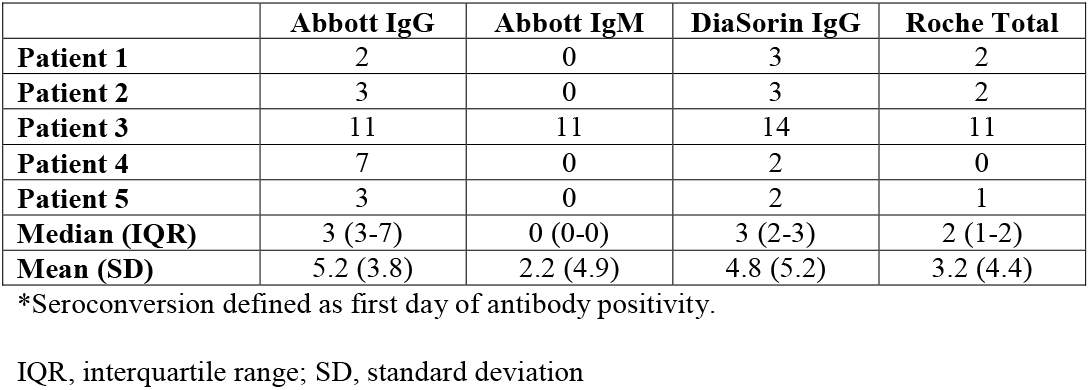
Days from PCR positivity to seroconversion

### Positive predictive value and negative predictive value

PPV and NPV for the four serologic assays across different seroprevalence estimates (1%, 5%, 10%) <14 days post-PCR positivity and ≥14 days are shown in **Table 5**. PPV for all four serologic assays was 100% across all seroprevalence estimates due to the specificity of 100% determined in this study. The NPV across all assays was higher for samples collected ≥14 days post-PCR positivity compared to <14 days, ranging from 97.7%-99.8% and 95.4%-99.7%, respectively. Notably, NPV decreased with increasing seroprevalence.

**Table 5.**
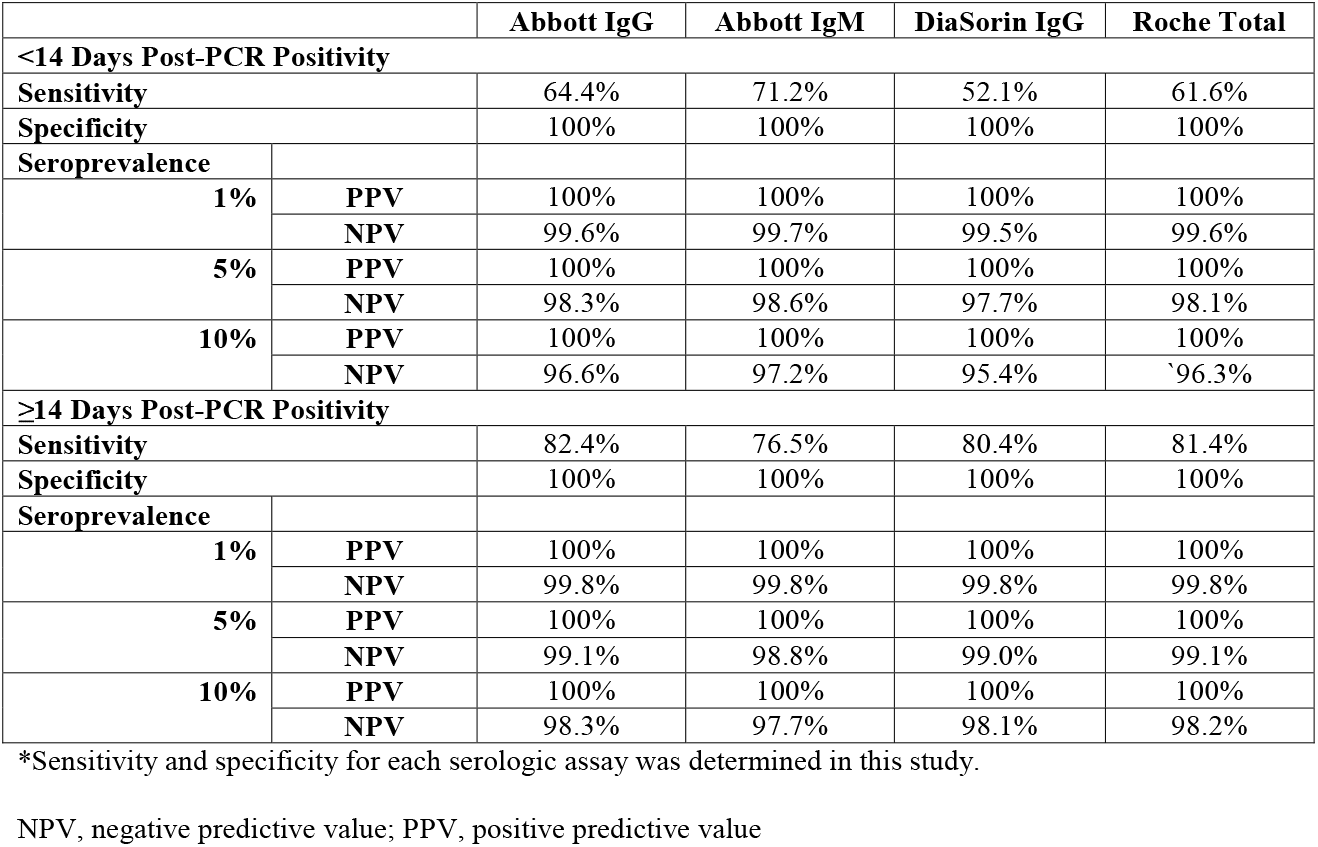
Positive and negative predictive values at 1%, 5%, and 10% seroprevalence for four anti-SARS-CoV-2 serologic assays

|  |  | Abbott IgG | Abbott IgM | DiaSorin IgG | Roche Total |
| --- | --- | --- | --- | --- | --- |
| **<14 Days Post-PCR Positivity** |  |  |  |  |  |
| **Sensitivity** |  | 64.4% | 71.2% | 52.1% | 61.6% |
| **Specificity** |  | 100% | 100% | 100% | 100% |
| **Seroprevalence** |  |  |  |  |  |
| **1%** | **PPV** | 100% | 100% | 100% | 100% |
|  | **NPV** | 99.6% | 99.7% | 99.5% | 99.6% |
| **5%** | **PPV** | 100% | 100% | 100% | 100% |
|  | **NPV** | 98.3% | 98.6% | 97.7% | 98.1% |
| **10%** | **PPV** | 100% | 100% | 100% | 100% |
|  | **NPV** | 96.6% | 97.2% | 95.4% | 96.3% |
| **≥14 Days Post-PCR Positivity** |  |  |  |  |  |
| **Sensitivity** |  | 82.4% | 76.5% | 80.4% | 81.4% |
| **Specificity** |  | 100% | 100% | 100% | 100% |
| **Seroprevalence** |  |  |  |  |  |
| **1%** | **PPV** | 100% | 100% | 100% | 100% |
|  | **NPV** | 99.8% | 99.8% | 99.8% | 99.8% |
| **5%** | **PPV** | 100% | 100% | 100% | 100% |
|  | **NPV** | 99.1% | 98.8% | 99.0% | 99.1% |
| **10%** | **PPV** | 100% | 100% | 100% | 100% |
|  | **NPV** | 98.3% | 97.7% | 98.1% | 98.2% |

\*Sensitivity and specificity for each serologic assay was determined in this study.
NPV, negative predictive value; PPV, positive predictive value

## Discussion

Here, we report analytical and clinical performance characteristics of the Abbott anti-SARS-CoV-2 IgM assay, which is not yet Health Canada approved but has received FDA EUA, alongside three other serologic assays: Abbott anti-SARS-CoV-2 IgG, DiaSorin anti-SARS-CoV-2 IgG, and Roche anti-SARS-CoV-2 Total. For the Abbott anti-SARS-CoV-2 IgM assay, sensitivity was 71.2% and 76.5% for <14 days and ≥14 days post-PCR positivity, respectively, with the highest sensitivity (87.5%) between 15-30 days post-PCR positivity. Combining Abbott anti-SARS-CoV-2 IgM and IgG testing improved sensitivity by 22.7% compared to anti-SARS-CoV-2 IgG testing alone, when tested the same day as PCR positivity. Specificity was 100% for all four serologic assays and only Abbott anti-SARS-CoV-2 IgG exhibited cross-reactivity for a sample positive for anti-dsDNA. Trends in anti-SARS-CoV-2 IgM over time since PCR positivity revealed heterogeneity across five patients, although confirming earlier IgM positivity compared to IgG in most patients.

Chemiluminescent anti-SARS-CoV-2 serologic assays, as used in this study, have been reported to exhibit high sensitivity, as summarized in a systematic review and meta-analysis (21). The highest pooled sensitivity for anti-SARS-CoV-2 serologic assays was observed for chemiluminescent assays (i.e. 97.8%), followed by enzyme-linked immunosorbent assays (84.3%) and lateral flow immunoassays (66.0%) (21), the latter most commonly used in point-of-care testing systems. Furthermore, anti-SARS-CoV-2 antibodies were reported to have higher pooled sensitivity at least three weeks after symptom onset compared to the first week and there was no association between immunoglobulin class and pooled sensitivity or specificity (21). The higher overall pooled sensitivity reported in this systematic review and meta-analysis compared to our study is likely due to its higher combined sample size of over 29,000 tests and the fact that 60% of our samples were analyzed within three weeks of PCR positivity. Perkmann, et al. also evaluated Abbott anti-SARS-CoV-2 IgG, DiaSorin anti-SARS-CoV-2 IgG, and Roche anti-SARS-CoV-2 total and reported sensitivities more similar to those observed in our study, ranging from 83.1% to 89.2% in 65 samples from COVID-19 patients evaluated ≥14 days after symptom onset (22). However, as is apparent in our study and others (10,23), sensitivity of the different immunoglobulin classes differ depending on days post symptom onset or PCR positivity.

As IgM is a marker of acute infection, anti-SARS-CoV-2 IgM sensitivity has been reported to be higher between 8-14 days (94.4%) compared to ≥15 days (89.5%), likely reflecting antibody class switching (24). While PCR sensitivity was higher than anti-SARS-CoV-2 IgM ELISA before 5.5 days post-symptom onset, IgM sensitivity was higher than PCR after 5.5 days post-symptom onset (10). Overall, sensitivity increased from 51.9% to 98.6% when testing PCR-negative patients with IgM ELISA compared to PCR testing alone (10), supporting its potential use as an adjunct test for early diagnosis of COVID-19. While we did not compare the performance of PCR and IgM testing, we did report serologic positivity the same day as positive molecular-based SARS-CoV-2 RNA detection, supporting its ability to detect antibody response while viral shedding is still occurring. Furthermore, we found that measuring anti-SARS-CoV-2 IgM and IgG on the same sample exhibits greater sensitivity than anti-SARS-CoV-2 IgG testing alone, especially when measured on the same day as PCR positivity. While anti-SARS-CoV-2 IgG assays exhibit good clinical performance, their greatest utility is seen two weeks after symptom onset (23,25–27). For example, sensitivity of the Abbott SARS-CoV-2 IgG assay was reported to reach 100% at day 17 after symptom onset (day 13 after PCR positivity) when assessing 689 serum specimens (25) and to exhibit an overall sensitivity of 78.3%, reaching 100% sensitivity 15 days after symptom onset when assessing 141 serum specimens (26). We report a similar trend with Abbott SARS-CoV-2 IgG sensitivity 64.4% <14 days post-PCR positivity and 82.4% ≥14 days and DiaSorin anti-SARS-CoV-2 IgG sensitivity 52.1% <14 days and 80.4% ≥14 days.

Our study reported 100% specificity for all four serologic assays using 52 samples collected pre-COVID-19 or from PCR negative patients. Previous studies reported slightly lower specificities with higher sample sizes. A meta-analysis reported overall specificity for anti-SARS-CoV-2 IgG and IgM assays of 99.1% (44 studies included) and 98.7% (41 studies included), respectively (9). Perkmann, et al. previously reported specificities of 99.2% for Abbott anti-SARS-CoV-2 IgG, 98.3% for DiaSorin anti-SARS-CoV-2 IgG, and 99.7% for Roche anti-SARS-CoV-2 total using over 1,154 pre-COVID-19 specimens (22). Other studies reported specificities of 99.9% using 1,020 serum specimens collected prior to SARS-CoV-2 circulation (25) and 99.3% using 107 serum specimens collected either before the emergence of SARS-CoV-2 or from PCR negative patients (26) for Abbott anti-SARS-CoV-2 IgG.

The trend observed in anti-SARS-CoV-2 IgG (Abbott) index values measured in serial samples over time were previously shown to vary among eight COVID-19 patients confirmed with PCR positivity (25). Some patients reached the positivity threshold by 12 days post-symptom onset, others plateaued after 12-16 days, while others continued to rise after 16 days (25). One study of sixty-three patients with confirmed COVID-19 collected samples at 3-day intervals until discharge and reported seroconversion in 96.8% of patients with the median time to seroconversion of 13 days (12). Interestingly, three seroconversion types were observed: synchronous IgG and IgM seroconversion, IgM seroconversion earlier than IgG, or IgM seroconversion later than IgG (12). The trends observed in our study were similarly heterogeneous between patients and the average time to seroconversion was 5.2, 2.2, 4.8, and 3.2 days post-PCR positivity for Abbott anti-SARS-CoV-2 IgG, Abbott anti-SARS-CoV-2 IgM, DiaSorin anti-SARS-CoV-2 IgG, and Roche anti-SARS-CoV-2 total, respectively.

Our study is not without limitations. Firstly, we did not collect information on the day of symptom onset due to confidentiality of personal health information, and thus all temporal data is only depicted as days since PCR positivity. This limits the accuracy of our assessment of sensitivity and antibody trends, as they are dependent on when the individual was tested for SARS-CoV-2 infection by PCR. However, this measure does reduce the potential subjectivity of using day of symptom onset as defined by each individual patient. Furthermore, as we did not collect information on clinical presentation and/or symptom severity, we are unable to make correlations between the assay signal and/or duration of antibody positivity with disease severity and/or patient outcome.

In conclusion, we report a sensitivity of 87.5% 15-30 days post-PCR positivity, an overall specificity of 100%, and no cross-reactivity with patient samples containing autoantibodies, viral antigens, or viral antibodies for the Abbott anti-SARS-CoV-2 IgM assay. Combining Abbott anti-SARS-CoV-2 IgM and IgG testing improved sensitivity by 22.7% compared to IgG testing alone when tested the same day as PCR positivity. The Abbott anti-SARS-CoV-2 IgM assay exhibited the earliest response and greatest signal in the majority of patients evaluated with serial sampling and had the highest NPV <14 days post-PCR positivity, suggesting its potential utility as an adjunct test to PCR early in disease course.

## Data Availability

The authors agree to make data available upon request.

## Acknowledgements

We would like to thank the UHN Core Lab Specimen Management team, Tech IVs, and biochemists for their help with sample retrieval and input. VK serves as a consultant for Abbott Diagnostics. DD and MB are employed by Abbott. Reagent kits were provided by Abbott Diagnostics. This research received no specific grant from any funding agency in the public, commercial, or not-for-profit sectors.

