## Supplementary material for "Analytical and clinical evaluation of four anti-SARS-CoV-2 serologic (IgM, IgG, and total) immunoassays": Table S1, Fig. S1, Fig. S2

**Table S1.** Method details and performance characteristics of four anti-SARS-CoV-2 serologic assays.

| | **Abbott ARCHITECT® SARS-CoV-2 IgG** | **Abbott ARCHITECT® SARS-CoV-2 IgM** | **DiaSorin LIAISON® SARS-CoV-2 S1/S2 IgG** | **Roche Elecsys® Anti-SARS-CoV-2** |
|---|---|---|---|---|
| **Antigen** | Nucleocapsid | RBD of the spike protein (S1 subunit) | Spike protein (S1 and S2 subunits) | Nucleocapsid |
| **Antibodies detected** | IgG | IgM | IgG | Total antibodies (IgA, IgG, IgM) |
| **Detection method** | CMIA | CMIA | CLIA | ECLIA |
| **Positivity cut-off** | ≥1.4 Index (S/C) | ≥1.00 Index (S/C) | ≥15.0 AU/mL | ≥ 1.0 COI |
| **Sensitivity** | <3 days post-symptom onset (n=5): 0.0% (0.0-52.2%)<br>3-7 days post-symptom onset (n=10): 50.0% (18.7-81.3%)<br>8-13 days post-symptom onset (n=34): 91.2% (76.3-98.1%)<br>≥14 days post-symptom onset (n=73): 100% (95.1-100%) | ≤7 days post-PCR (n=158): 59.5% (51.7-66.8%)<br>8-14 days post-PCR (n=109): 94.5% (88.5-97.5%)<br>15-30 days post-PCR (n=55): 100% (93.5-100%)<br>≥31 days post-PCR (n=4): 100% (51.0-100%) | ≤5 days from diagnosis (n=44): 25.0% (14.6-39.4%)<br>6-14 days from diagnosis (n=18): 89.8% (78.2-95.6%)<br>>15 days from diagnosis (n=14): 97.6% (87.4-99.6%) | 0-6 days post-PCR (n=161): 60.2% (52.3-67.8%)<br>7-13 days post-PCR (n=150): 85.3% (78.6-90.6%)<br>≥14 days post-PCR (n=185): 99.5% (97.0-100%) |
| **Specificity** | Overall (n=1070): 99.6% (99.1-99.9%)<br>Pre-COVID-19 Outbreak (n=997): 99.6% (99.0-99.9%)<br>Other Respiratory Ilness (n=73): 100% (95.1-100%) | Overall (n=2965): 99.6% (99.3-99.7%) | Overall (n=1090): 99.3% (98.6-99.6%)<br>*All samples collected prior to COVID-19 | Overall (n=10453): 99.8% (99.7-99.9%)<br>Diagnostic routine (n=6305): 99.8% (99.7-99.9%)<br>Blood donors (n=4148): 99.8% (99.6-99.9%)<br>*All samples obtained before December 2019 |
| **Cross-Reactivity** | Overall agreement 99.5% (181/182) when testing specimens containing antibodies against several viruses, known autoimmune disorders, and other medical conditions.<br>Cross-reaction observed for: CMV (IgG+) | Overall agreement 99.0% (206/208) when testing specimens containing antibodies against several viruses, known autoimmune disorders, and other medical conditions.<br>Cross-reaction observed for: Hemodialysis patient and RF | Overall agreement 98.2% (165/168) when testing specimens containing antibodies against several viruses, known autoimmune disorders, and other medical conditions.<br>Cross-reaction observed for: Anti-HBV, anti-influenza A, RF | Overall agreement 99.5% (788/792) when testing specimens containing antibodies against several viruses, known autoimmune disorders, other human coronaviruses.<br>Cross-reaction observed for: EBV acute (IgM+, IgG+), CMV acute (IgM+, IgG+), SLE |

CLIA, chemiluminescent immunoassay; CMIA, chemiluminescent microparticle immunoassay; CMV, cytomegalocirus; COI, cutoff index; EBV, Epstein-Barr virus; ECLIA, electrochemiluminescence immunoassay; HBV, hepatitis B virus; RBD, receptor-binding domain; RF, rheumatoid factor; SLE, systemic lupus erythematosis; NPA, negative percent agreement; PCR, polymerase chain reaction; PPA, positive percent agreement

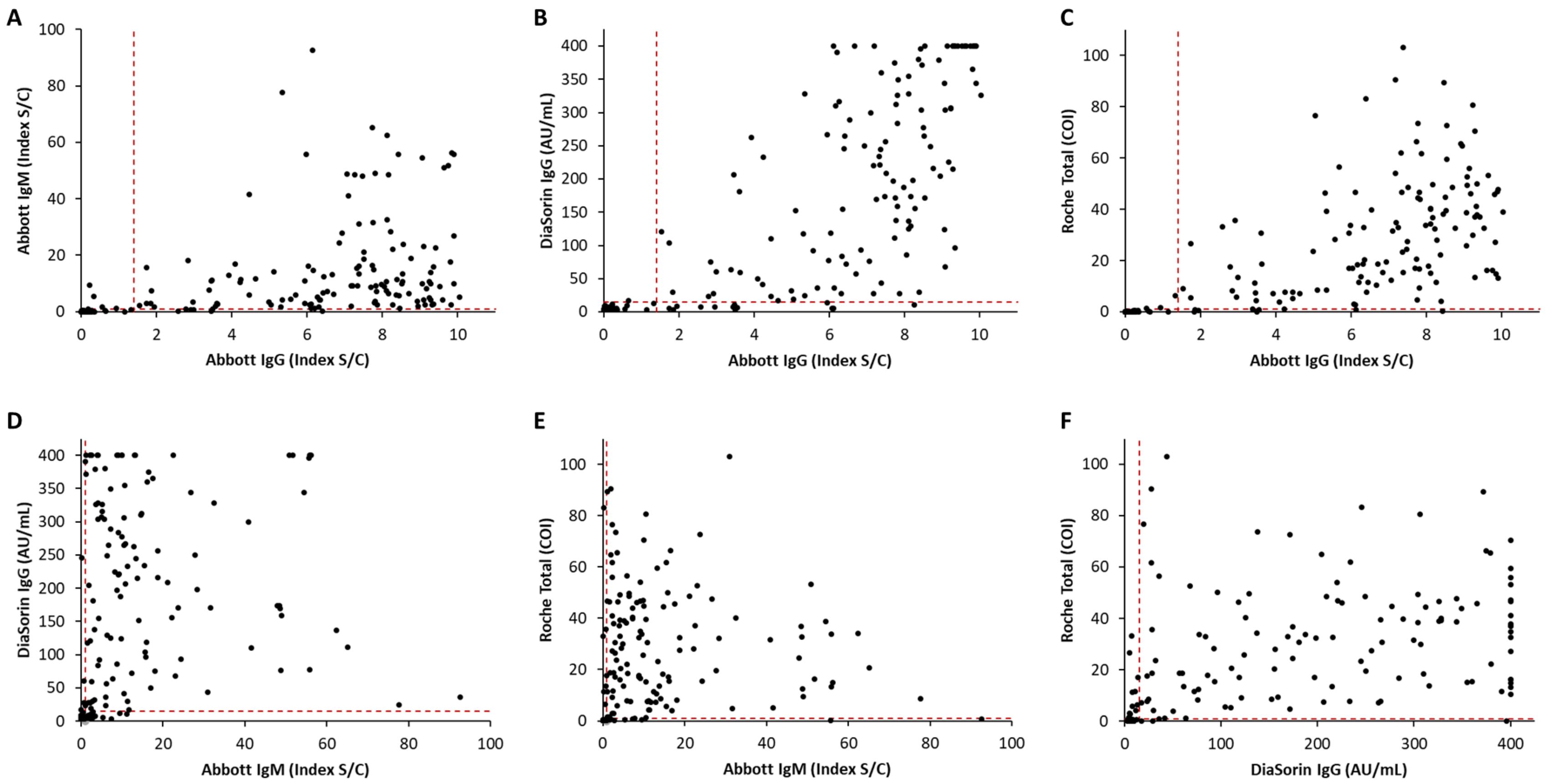


**Fig. S1**. **Concordance between four anti-SARS-CoV-2 serologic immunoassays.** Anti-SARS-CoV-2 positivity or negativity for 280-282 serum and plasma patient samples as determined by four serologic immunoassays is shown. 175 patients were PCR positive for SARS-CoV-2. Dotted lines represent positivity cut-offs for each assay.

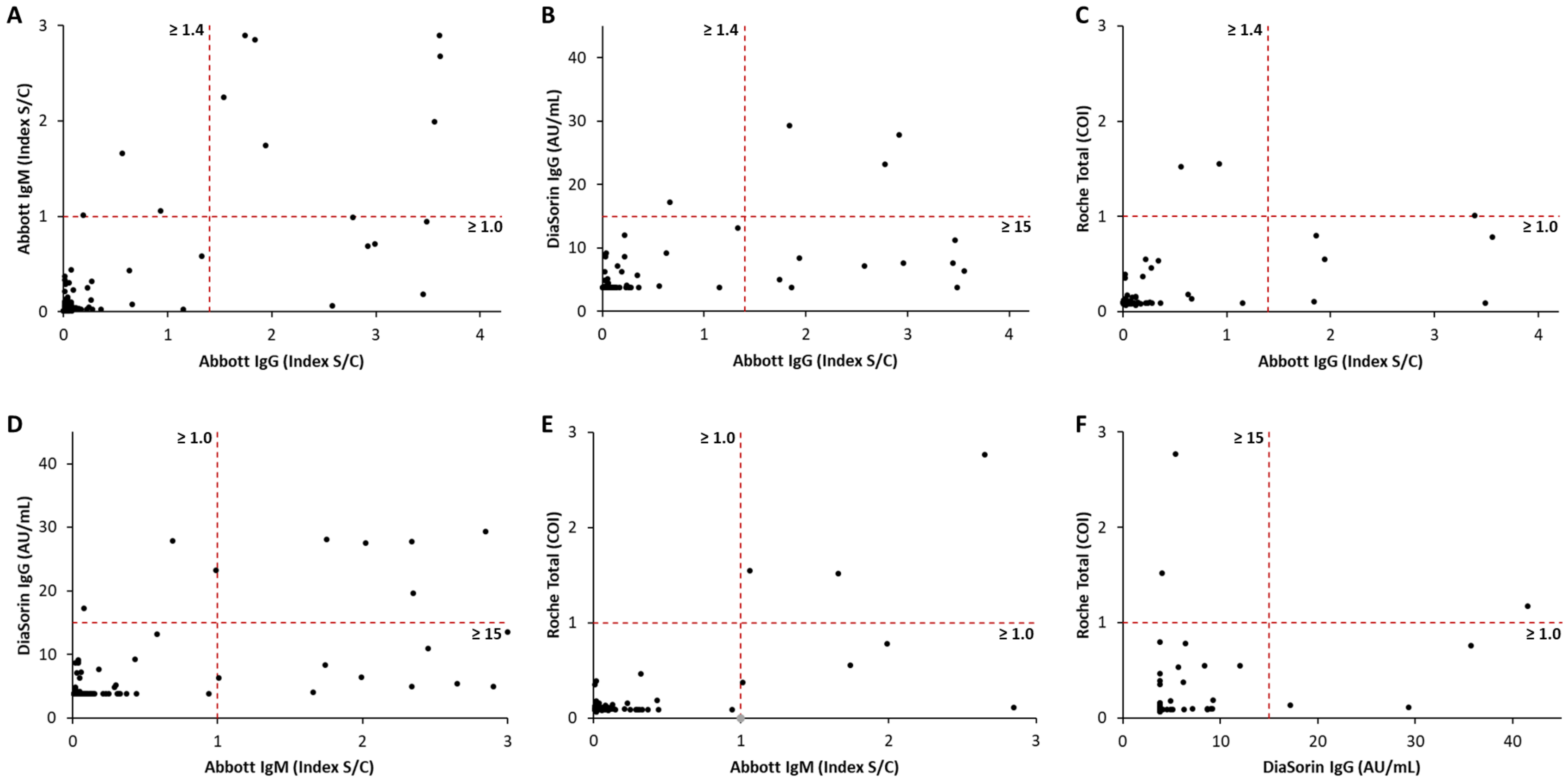


**Fig. S2. Concordance between four anti-SARS-CoV-2 serologic immunoassays (zoomed in).** Anti-SARS-CoV-2 positivity or negativity for 280-282 serum and plasma patient samples as determined by four serologic immunoassays is shown. 175 patients were PCR positive for SARS-CoV-2. Axes were shortened to 3 times the positivity cut-off to allow better visualization of concordance around the positivity cut-off. Dotted lines represent positivity cut-offs for each assay and each positivity cut-off is stated.
